# FALSE-NEGATIVE RESULTS OF INITIAL RT-PCR ASSAYS FOR COVID-19: A SYSTEMATIC REVIEW

**DOI:** 10.1101/2020.04.16.20066787

**Authors:** Ingrid Arevalo-Rodriguez, Diana Buitrago-Garcia, Daniel Simancas-Racines, Paula Zambrano-Achig, Rosa Del Campo, Agustín Ciapponi, Omar Sued, Laura Martínez-García, Anne Rutjes, Nicola Low, Patrick M. Bossuyt, Jose A Perez-Molina, Javier Zamora

## Abstract

**Background:** A false-negative case of severe acute respiratory syndrome coronavirus 2 (SARS-CoV- 2) infection is defined as a person with suspected infection and an initial negative result by reverse transcription-polymerase chain reaction (RT-PCR) test, with a positive result on a subsequent test. False-negative cases have important implications for isolation and risk of transmission of infected people and for the management of coronavirus disease 2019 (COVID-19). We aimed to review and critically appraise evidence about the rate of RT-PCR false-negatives at initial testing for COVID-19.

**Methods:** We searched MEDLINE, EMBASE, LILACS, as well as COVID-19 repositories including the EPPI-Centre living systematic map of evidence about COVID-19 and the Coronavirus Open Access Project living evidence database. Two authors independently screened and selected studies according to the eligibility criteria and collected data from the included studies. The risk of bias was assessed using the Quality Assessment of Diagnostic Accuracy Studies (QUADAS-2) tool. We calculated the proportion of false-negative test results with the corresponding 95% CI using a multilevel mixed-effect logistic regression model. The certainty of the evidence about false-negative cases was rated using the GRADE approach for tests and strategies. All information in this article is current up to July 17, 2020.

**Results:** We included 34 studies enrolling 12,057 COVID-19 confirmed cases. All studies were affected by several risks of bias and applicability concerns. The pooled estimate of false-negative proportion was highly affected by unexplained heterogeneity (tau-squared= 1.39; 90% prediction interval from 0.02 to 0.54). The certainty of the evidence was judged as very low, due to the risk of bias, indirectness, and inconsistency issues.

**Conclusions:** There is a substantial and largely unexplained heterogeneity in the proportion of false-negative RT-PCR results. The collected evidence has several limitations, including risk of bias issues, high heterogeneity, and concerns about its applicability. Nonetheless, our findings reinforce the need for repeated testing in patients with suspicion of SARS-CoV-2 infection given that up to 54% of COVID-19 patients may have an initial false-negative RT-PCR (certainty of evidence: very low). An update of this review when additional studies become available is warranted.

**Systematic review registration:** Protocol available on the OSF website: https://osf.io/gp38w/

## INTRODUCTION

On December 31, 2019, the World Health Organization (WHO) was alerted about a cluster of patients with pneumonia in Wuhan City, Hubei province, China (1). Chinese authorities confirmed, a week later, an outbreak of a novel coronavirus. The virus has been named as severe acute respiratory coronavirus 2 (SARS-CoV-2) (SARS-CoV-2) (2), and the clinical disease that it causes is coronavirus disease 2019 (COVID-19), which has become a worldwide public health emergency and reached pandemic status (3). By the time of this article’s writing, the virus has spread to 215 countries and territories and has caused over 283,271 deaths worldwide (4).

Clinical suspicion of COVID-19 is based primarily on respiratory symptoms such as fever, cough, and shortness of breath as primary manifestations (5, 6). The spectrum of symptoms and clinical signs associated with COVID-19 has expanded with increasing knowledge about SARS-CoV-2. Although most of the cases present mild symptoms, some cases have developed pneumonia, severe respiratory diseases, kidney failure, and even death (7-9). SARS-CoV-2 mainly spreads through person-to-person contact via respiratory droplets from coughing and sneezing, and through surfaces that have been contaminated with these droplets (10). Recent evidence has suggested the presence of asymptomatic cases in several different settings showing, that the proportion could be up to 29% (11). Furthermore, recent studies have shown the presence of asymptomatic cases in cluster families, possibly transmitting the virus before a virus-carrying person displays any symptom (12-14).

Because the signs of infection mentioned above are non-specific, confirmation of cases is currently based on the detection of nucleic acid amplification tests that detect viral ribonucleic acid (RNA) sequences by reverse transcription-polymerase chain reaction (RT-PCR). Different RT-PCR assays have been proposed, all of which include the N gene that codes for the viral nucleocapsid. Other alternative targets are the E gene, for the viral envelope; the S gene for the spike protein; and the Hel gene for the RNA polymerase gene (RdRp/Helicase) (15, 16). Molecular criteria for *in vitro* diagnosis of COVID-19 disease are heterogeneous, and usually require the detection of two or more SARS-CoV-2 genes (17).

RT-PCR repeated testing might be required to confirm a clinical diagnosis, especially in the presence of symptoms closely related to COVID-19, as numerous clinical practice guidelines and consensus statements recommend (18-22). Cases with negative RT-PCR results at initial testing and found to be positive in a subsequent test are commonly considered cases with an initial false-negative diagnosis. Some researchers have suggested that these failures in SARS-CoV-2 detection are related to multiple preanalytical and analytical factors, such as lack of standardisation for specimen collection, delays or poor storage conditions before arrival in the laboratory, the use of inadequately validated assays, contamination during the procedure, insufficient viral specimens and load, the incubation period of the disease, and the presence of mutations that escape detection or PCR inhibitors (17, 23).

The availability of accurate laboratory tools for COVID-19 is essential for case identification, contact tracing, and optimisation of infection control measures, as it was shown by previous epidemics caused by SARS-CoV and the Middle East Respiratory Syndrome Coronavirus (MERS-CoV) (24-26). Due to the significant burden on health systems around the globe caused by the COVID-19 pandemic, and the potential consequences at several levels of missing a COVID-19 case, we aimed to obtain through a systematic review of the literature, a summary estimate of the proportion of false-negatives related to the detection of SARS-CoV-2 using RT-PCR assays at the first healthcare encounter (initial testing).

## MATERIALS AND METHODS

We followed the Preferred Reporting Items for Systematic Reviews and Meta-Analyses of Diagnostic Test Accuracy studies (PRISMA-DTA) to prepare this report (27). A protocol for this review, as well as previous reports of findings by date of search, are available in the Open Science Framework repository for public consultation (https://osf.io/iserd/).

### Criteria for considering studies for this review

We included observational studies (including accuracy studies) reporting the initial use of RT-PCR in the detection of SARS-CoV-2 RNA in patients under suspicion of infection by clinical or epidemiological criteria. We primarily aimed to include studies enrolling consecutive patients who were receiving RT-PCR at first healthcare encounter (initial testing), with further confirmation of SARS-CoV-2 infection and/or COVID-19 diagnosis (positive/negative) by an additional RT-PCR evaluation. We did not impose limits by age, gender, or study location.

We aimed to include all types of RT-PCR kits, regardless of the brand or manufacturer, the RNA extraction method used, the number of target gene assays assessed, or the cycle threshold value for positivity. We excluded studies focus on other populations or reporting samples/specimens instead of patients (such as monitoring or discharge of COVID-19 confirmed cases, population screening and patients with high-risk comorbidities), studies only providing the absolute number of false-negatives or without clear information about numerical information, as well as studies reporting validation of novel assays or comparing sample collection/sample specimens (i.e. focus on agreement). Full eligibility criteria can be found in the S1 Appendix.

### Search methods for identification of studies

We carried out a comprehensive and sensitive search strategy based on search terms developed for the COVID-19 Open Access Project by researchers and librarians at the Institute of Social and Preventive Medicine, University of Bern (https://ispmbern.github.io/covid-19/living-review/collectingdata.html) in the following databases:

- MEDLINE (Ovid SP, 1946 to July 17, 2020)
- Embase (Ovid SP, 1982 to July 17, 2020)
- LILACS (iAH English) (BIREME, 1982 to July 17, 2020)

We did not apply any language restrictions to electronic searches (S2 Appendix). As additional sources of potential studies, we searched in repositories of preprint articles, clinical trials registries for ongoing or recently completed trials (clinicaltrials.gov; the World Health Organization’s International Trials Registry and Platform, and the ISRCTN Registry), and the reference lists of all relevant papers. Finally, we also screened the following resources for additional information:

- The WHO Database of publications on coronavirus disease (COVID-19) (Available on https://www.who.int/emergencies/diseases/novel-coronavirus-2019/global-research-on-novel-coronavirus-2019-ncov).
- The LOVE (Living Overview of Evidence) centralised repository developed by Epistemonikos (available on https://app.iloveevidence.com/topics)
- The Living systematic map of the evidence about COVID-19 produced by EPPI-Centre (28).
- The COVID-19 Open Access Project Living Evidence on COVID-19, developed at the Institute of Social and Preventive Medicine, University of Bern (available on https://ispmbern.github.io/covid-19/)

### Data collection and analysis

For the selection of eligible studies, two reviewers independently screened the search results based on their titles and abstract. We retrieved the full-text copy of each study assessed as potentially eligible, and pairs of reviewers confirmed eligibility according to the selection criteria. In case of disagreements, we reached consensus by discussion. For data extraction, one reviewer extracted qualitative and quantitative data from eligible studies, and an additional reviewer checked all the extracted information for accuracy. We contacted study authors to supply missing information about critical characteristics of included studies.

### Assessment of methodological quality

Two authors independently assessed the risk of bias of included studies, and disagreements were resolved through discussion. We evaluated the methodological quality using the Quality Assessment of Diagnostic Accuracy Studies (QUADAS-2) tool (29). We decided to also apply the QUADAS-2 tool for case series studies due to the lack of tools to assess the risk of bias associated with these studies. However, for a more comprehensive assessment of the limitations of the included studies, we adapted the Joanna Briggs Institute Critical Appraisal Checklist for Case Series (30). This tool included items about inclusion criteria, measurement of asymptomatic status, follow-up of the course of the disease, and availability of numerator and denominator. We added questions about the representativeness of the source and target populations as well.

### Statistical analysis and data synthesis

For all included studies, we extracted data about the number of false-negative cases as well as the total of confirmed cases by additional RT-PCR investigations (i.e. repeated testing). We presented the results of estimated proportions (with 95% CIs) in a forest plot to assess the between-study variability. We aimed to calculate a summary estimate of the false-negative rate with the corresponding 95% CI using a multilevel mixed-effect logistic regression model in Stata 16^®^. This method allowed us to estimate the between-study heterogeneity from the variance of study-specific random intercepts. We computed 90% prediction intervals to include the between-study variation. The 90% prediction interval shows the range of true false-negative proportions that can be expected in 90% of future settings, comparable to the ones included in the meta-analysis. We expressed heterogeneity in primary study results using the Tau-square statistic.

We planned to investigate the potential sources of heterogeneity using a descriptive approach and performing a random-effects meta-regression analysis, including covariates, one at each time, into the logistic model. Anticipated sources of heterogeneity included the type of specimen collected, the presence or not of clinical findings, the number of RNA targets genes under assessment, and the time of symptom evolution.

### Summary of findings and certainty of the evidence

We rated the certainty of the evidence about false-negative cases following the GRADE approach for tests and strategies (31, 32). We assessed the quality of evidence as high, moderate, low, or very low, depending on several factors, including risk of bias, imprecision, inconsistency, indirectness, and publication bias. We illustrate the consequences of the numerical findings in a population of 100 tested, according to three different prevalence estimates of the disease provided by the stakeholders involved in this review.

## RESULTS

Electronic searches yielded 2536 references from the selected databases. In addition, we obtained 186 additional references searching in other resources (Figure 1). Our initial screening of titles and abstracts identified 171 references to assess in full text. We excluded 137 studies mostly due to: a) ineligible setting (no initial COVID-19 testing); b) incomplete or no data about false-negative cases and COVID-19 confirmed cases; c) ineligible population (i.e. pooling sample, analysis based on samples instead of patients) (S3 Appendix). We included 34 studies in our synthesis (33-66) which dealt with 12057 patients (Table 1).

**Figure 1.**
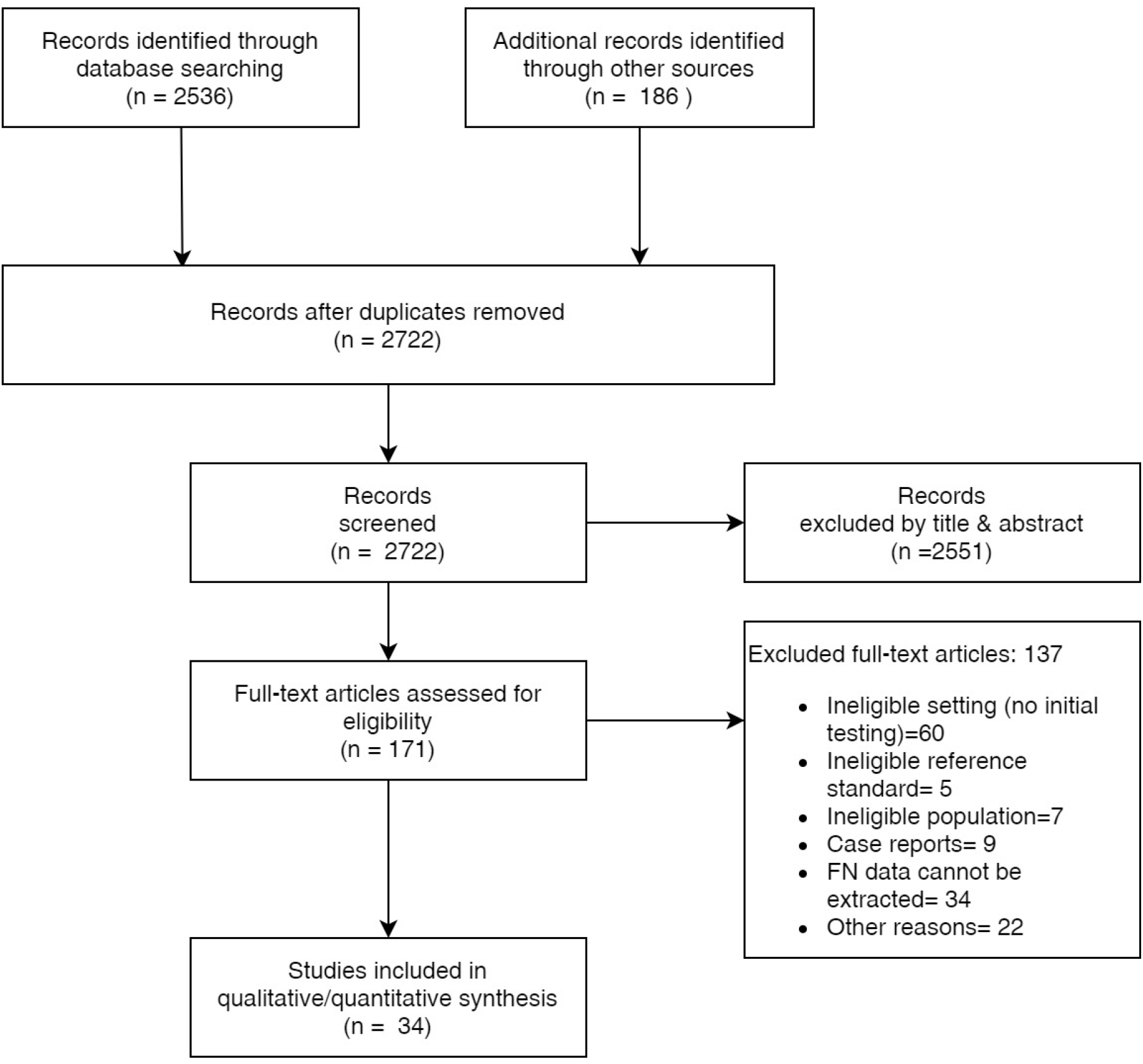
PRISMA flow diagram.

**Table 1.** Characteristics of included studies.

| ID | Data collection | Country | Setting | Age (years) | % Male: % Female | Type of specimen | RT-PCR Brand | Target genes | Days from symptoms onset (days) |
| --- | --- | --- | --- | --- | --- | --- | --- | --- | --- |
| Ai T 2020 (33) | January 6-February 6 | China | Tongji Hospital of Tongji Medical College of Huazhong University of Science and Technology, Wuhan, Hubei, China | Mean 51 ± 15<br>Range from 2 to 95<sup>b</sup> | 46:54<sup>b</sup> | Throat swab | TaqMan One-Step RT-PCR Kits from Shanghai Huirui Biotechnology Co., Ltd<br>Shanghai BioGerm Medical Biotechnology Co., Ltd | Not reported | Not reported |
| Albert E 2020 (34) | Unclear-April 14 | Spain | Hospital Clínico Universitario of Valencia | Median 65 years; range from 3 to 98<sup>c</sup> | 57:43<sup>c</sup> | Nasopharyngeal or oropharyngeal swabs, upper RT samples | LightMix Modular SARS-CoV-2 (COVID-19) E-gene/LightMix Modular SARS-CoV-2 (COVID-19) RdRP gene from TIB MOLBIOL GmHD<br>SARS-CoV-2 Real-time PCR Kit from Vircell Diagnostics<br>SARS-CoV-2 (S gene)-BD Max System (Viasure Real-Time PCR Detection Kits; CerTest, Zaragoza, Spain). | E, RdRp, S | Median 5 days; range: 1-14 days |
| Bernheim A 2020 (35) | January 18-February 2 | China | Hospitals from four provinces in China: Nanchang (Jiangxi Province), Zhuhai (Guangdong Province), Chengdu (Sichuan province) and Guilin (Guangxi province) | Mean 45 ± 15,6<sup>b</sup> | 50:50<sup>b</sup> | Bronchoalveolar lavage, endotracheal aspirate, nasopharyngeal swab, or oropharyngeal swab | Sansure Biotech Inc. (Changsha, China), Shanghai Zhijiang Biotechnology Co. (Shanghai, China),<br>Da An Gene Co. (Guangzhou, China). | Not reported | Range from 0 to 12 |
| Besutti G 2020 (36) | March 13-23 | Italy | AUSL-IRCCS di Reggio Emilia, Reggio Emilia, Italy | Mean 59 ± 15.8<sup>b</sup> | 59:41<sup>b</sup> | Nasopharyngeal and oropharyngeal swabs | GeneFinder™ COVID -19 PLUS Real Real Amp Kit | Not reported | Not reported |
| Chen D 2020 (37) | January 19-February 20 | China | Five non-specialised infectious disease hospitals in Guangzhou | Mean 49.7 ± 15.7<sup>a</sup> | 43:57<sup>a</sup> | Nasopharyngeal or oropharyngeal swabs | Not reported | Not reported | Not reported |
| Chen HJ 2020 (38) | January 26-February 4 | China | Hainan General Hospital | Mean 54.5 ± 11.8<sup>a</sup> | 68:32<sup>a</sup> | Not reported | Not reported | Not reported | Mean 6,3 ± 5,6 days |
| Chen ZH 2020 (39) | January 24-February 6 | China | The Hangzhou Xixi Hospital Affiliated to Zhejiang Chinese Medical University | Mean 46.9 ± 11.1<sup>a</sup> | 55:45<sup>a</sup> | Not reported | Not reported | Not reported | Mean 2; range 1 to 4,5 days |
| Çinkoğlu A 2020 | March 15-April 15 | Turkey | Ege University Faculty of Medicine | Means from 39.9 to 51<sup>a</sup> | 47:53<sup>a</sup> | Not reported | Not reported | Not reported | Not reported |
| (40) |  |  |  |  |  |  |  |  |  |
| Dai H 2020 (41) | January 10-February 7 | China | 13 hospitals in Jiangsu | Mean 44.6 ± 14.8<sup>a</sup> | 58:42<sup>a</sup> | Respiratory samples | Not reported | Not reported | Not reported |
| Duan X 2020 (42) | January 10-February 8 | China | The First Affiliated Hospital, College of Clinical Medicine, Medical College of Henan University of Science and Technology, Luoyang | Mean 52 ± 19.3<sup>a</sup> | 60:40<sup>a</sup> | Nasal and pharyngeal swab specimens | Not reported | Not reported | Mean 6,64 ± 3,82 days |
| Fang Y 2020 (43) | January 19-February 4 | China | Taizhou Enze Medical Center (Group) Enze Hospital, China | Median 45; IQR: 39-55<sup>a</sup> | 57:43<sup>a</sup> | Throat swab, sputum | Not reported | Not reported | Mean 3±3 |
| Fechner C 2020 (44) | Unclear | Switzerland | Cantonal Hospital Lucerne | Mean 63 ± 15.7<sup>a</sup> | 75:25<sup>a</sup> | Nasopharyngeal or oropharyngeal swabs | Not reported | Not reported | Not reported |
| Gietema 2020 (45) | March 13-24 | Netherlands | Maastricht University Medical Centre (MUMC+), the Netherlands | Median 66; IQR: 55-76<sup>b</sup> | 59:41<sup>b</sup> | Nasopharyngeal and/or oropharyngeal swab | Tib-Molbiol (Berlin, Germany)<br>Biolegio (Netherlands) | RdRp, E | Not reported |
| He JL 2020 (67) | January 10 – February 28 | China | University of Hong Kong-Shenzhen Hospital, China | Median 52; range: 8 to 74<sup>a</sup> | 50:50<sup>a</sup> | Nasopharyngeal swab, oropharyngeal swab, endotracheal aspirate, or bronchoalveolar lavage | BGI Genomics (Shenzhen, China) | Not reported | Not reported |
| Lan FY 2020 (47) | March 9-April 15 | USA | Massachusetts community healthcare system | Mean 43.6 ± 12.9<sup>b</sup> | 21:79<sup>b</sup> | Nasopharyngeal swabs | CDC 2019-Novel RT-PCR<br>Roche Cobas SARS-CoV-2<br>Abbott Real Time SARS-CoV-2 | Not reported | Not reported |
| Lee TH 2020 (48) | January-February 29 | Singapore | National Centre for Infectious Diseases, Singapore | Not reported | Not reported | Nasopharyngeal swabs, sputum, and stool if diarrhoea is present | Laboratory developed test<br>A*STAR Fortitude Kit (Accelerate Technologies, Singapore) | N +ORF1ab | Median 5 days; range from 1 to 24 days |
| Li Y 2020 (49) | February 2-17 | China | Hankou Hospital of Wuhan, China | Median 57; range: 22 to 88<sup>b</sup> | 56:44<sup>b</sup> | Pharyngeal swab specimens | Shengxiang Biotechnology Co (novel coronavirus 2019-nCoV nucleic acid detection kit (fluorescence PCR method)<sup>d</sup> | ORF1ab<sup>d</sup> | Not reported |
| Long C 2020 (50) | January 20-February 8 | China | Yichang Yiling Hospital, China | Mean 44,8 ± 18,2<sup>a</sup> | 56:44<sup>a</sup> | Not reported | DAAN GENE<sup>d</sup> | ORF1ab<sup>d</sup> | Only duration of fever reported: 2,6 ± 1,7 days |
| Long DR 2020 (51) | March 2-30 | USA | University of Washington Virology clinical laboratory | Means from 56.7 to 61.6<sup>c</sup> | 57:43<sup>c</sup> | Nasopharyngeal swabs | Laboratory-developed test (LDT) two-target/two-control assay modified from the CDC Panther Fusion SARS-CoV-2 assay (Hologic, Marlborough, MA, target genes two conserved | N1, N2, ORF1ab, E, S | Not reported |
|  |  |  |  |  |  |  | regions of ORF1ab);<br>Roche RT-PCR (Basel, Switzerland, target E gene)<br>DiaSorin (Saluggia, Italy, target ORF1ab and S genes). |  |  |
| Ma H 2020 (52) | January 21-February 14 | China | Wuhan Children's Hospital | Mean 2.5; range: 0.9 to 7<sup>a</sup> | 56:44<sup>a</sup> | Not reported | Not reported | Not reported | Not reported |
| Richards on 2020 (53) | March 1-April 4 | USA | 12 hospitals in New York City, Long Island, and Westchester County, New York (Northwell Health system), USA | Median 63; IQR: 52-75<sup>a</sup> | 60:40<sup>a</sup> | Nasopharyngeal swabs | Not reported | Not reported | Not reported |
| Shen N 2020 (54) | January 22-February 18 | China | Tongji Hospital in Wuhan | Median 56; IQR: 42-66 | 49:51 | Throat swabs | SARS-CoV-2 nucleic acid detection kit (Shanghai Huirui Biotechnology Co. Ltd) | N +ORF1ab | Not reported |
| Wang P 2020 (55) | January 25-March 16 | China | First People's Hospital of Jingmen, Hubei Province | Median 58; range: 21-95 | 46:54 | Throat swabs | RT-PCR reagent BioGerm (Shanghai BioGerm Medical Technology Co., Ltd.) | N +ORF1ab | Not reported |
| Wen Z 2020 (56) | January 21-February 14 | China | Two areas in Henan Province, China | Median 16; range: 12 to 98<sup>b</sup> | 47:53<sup>b</sup> | throat-swab, sputum, or alveolar lavage fluid specimens | Not reported | Not reported | Not reported |
| Wong HYF 2020 (57) | January 1-March 5 | China | Four tertiary and regional hospitals in Hong Kong (Queen Mary Hospital, Pamela Youde Nethersole Eastern Hospital, Queen Elizabeth Hospital, and Ruttonjee Hospital), China | Mean 56; range: 16 to 96<sup>a</sup> | 41:59<sup>a</sup> | nasopharyngeal swabs and throat swabs | QuantiNova Probe RT-PCR Kit (QIAGEN, Hilden, Germany) | RdRp | Not reported |
| Wu J 2020 (58) | January 22-February 14 | China | First People's Hospital of Yancheng City, the Second People's Hospital of Yancheng City, and the Fifth People's Hospital of Wuxi, China | Median 46.1; IQR: 30.7 to 61.5 | 49:51 | nose swab and/or throat swab | Bio-germ, Shanghai, China | N +ORF1ab | Not reported |
| Xie X 2020 (59) | January 16-February 2 | China | Database of Radiology Quality Control Centre, Hunan/ 3 cities in Hunan Province, China | Not reported | Not reported | swab test; no further details provided | Not reported | Not reported | Not reported |
| Young BE 2020 (60) | January 23-February 3 | Singapore | Four hospitals in Singapore | Median 47; range: 31-73<sup>a</sup> | 50:50<sup>a</sup> | Nasopharyngeal swabs | QuantiTect Probe RT-PCR kit (Qiagen) | N, S, ORF1ab | Median 13; range 5-24 days |
| Zhang H 2020 (61) | January 22-February 28 | China | Huanggang Central Hospital and The Second Affiliated Hospital of Shandong First Medical University | Median 48.3; IQR: 33-56<sup>a</sup> | 56:44<sup>a</sup> | Not reported | The Beijing Genomics Institute (BGI, Beijing, China) | Not reported | Not reported |
| Zhang JJ 2020 (62) | December 29-February 16 | China | Zhongnan Hospital of Wuhan University and No.7 hospital of Wuhan, China | Median 57; range: 22 to 88<sup>a</sup> | 53:47<sup>a</sup> | Pharyngeal swab | Shanghai bio-germ Medical Technology Co Ltd | N +ORF1ab | Not reported |
| Zhao JJ 2020 (63) | January 11-February 9 | China | Shenzhen Third People's Hospital | Median 48; IQR: 35-61<sup>a</sup> | 49:51<sup>a</sup> | Throat swabs, Nasal swabs | Not reported | Not reported | Not reported |
| Zhifeng 2020 (64) | January 25-February 6 | China | Xiaogan Central Hospital, China | Range: 23 to 82<sup>a</sup> | 59:41<sup>a</sup> | Throat swabs | Multiple brands<sup>d</sup> | N +ORF1ab | Mean 6,5 days<sup>d</sup> |
| Zhou H 2020 (65) | January 19-February 15 | China | First Affiliated Hospital, Zhejiang University School of Medicine | Mean 53.3; range: 14-96<sup>c</sup> | 59:41<sup>c</sup> | Bronchoalveolar lavage, endotracheal aspirate, or nasopharyngeal swab | Not reported | Not reported | Not reported |
| Zhou S 2020 (66) | January 16-February 12 | China | Tongji Hospital of Tongji Medical College, Huazhong University of Science and Technology | Mean 52.3 ± 13.1<sup>a</sup> | 54:46<sup>a</sup> | Pharyngeal swab | Not reported | Not reported | Not reported |

**Notes:** a) Information from COVID-19 confirmed cases only; b) Information from COVID-19 suspected (positive and negative); c) information from other groups reported by the authors; d) data provided by the corresponding author (personal communication).

The sample sizes ranged from 18 to 5,700 COVID-19 confirmed cases (median 90; interquartile range -IQR= 46.5 to 204). Twelve studies focused on the estimation of diagnostic test accuracy, including populations with suspected COVID-19 at the beginning of the study (33, 36-38, 40, 43, 45, 46, 50, 56, 64). The remaining studies reported information from case series, most of which included confirmed cases of COVID-19 at the beginning of the study (34, 35, 41, 42, 44, 47-49, 51-55, 57-63, 65, 66). One study focused its data collection only on children (52) and other only on healthcare workers (47). Only three studies included a small number of patients without symptoms at the time of testing (from two to nine patients), but they did not provide subgroup information of these cases (47, 52, 57).

Included studies collected information from January 1 (57) to April 15, 2020 (40, 47); two studies did not provide complete information about the time of recruitment (34, 44). Ten studies were carried out in institutions outside of China (34, 36, 40, 44, 45, 47, 48, 51, 53, 60). The age of participants was reported heterogeneously in 21 studies providing information of COVID-19 confirmed cases (37-44, 46, 50, 52-54, 57, 58, 60-64, 66): for studies reporting a mean, the average ranged from 2.5 (52) to 56 years (57), while for studies reporting medians, the corresponding range was 45 (43) to 63 years (53). These 21 studies reported a total of 5331 men and 4067 women (Table 1).

In all cases, the presence of infection was confirmed after detection of SARS-CoV-2 RNA in any real-time RT-PCR assay that was repeated after a negative result. The specimens collected for the RT-PCR assessment were heterogeneous in most of the included studies; in 13 studies the authors reported the use of nasopharyngeal swabs (34-37, 44-46, 48, 51, 53, 57, 60, 65), along with oropharyngeal swabs in 7 out of these 13 studies (34-37, 44-46) (Table 1). The name/brand of the SARS-CoV-2 nucleic acid detection kit used was reported by 19 studies (33-36, 45-51, 54, 55, 57, 58, 60-62, 64), and 13 studies reported the target genes under assessment for positivity (34, 45, 48-51, 54, 55, 57, 58, 60, 62, 64), with the ORF1ab being the most frequently used (8 studies). Ten studies provided heterogeneous information about the time from the symptom onset to initial testing (34, 35, 38, 39, 42, 43, 48, 50, 60, 64) (Table 1).

### Quality of included studies

We applied the QUADAS-2 tool to all included studies to reflect critical limitations in the validity of the findings (Figure 2). In addition, given that to some of the studies were cohorts/case series, we also applied the JBI tool for case series to all included studies for a comprehensive assessment of their limitations (S4 Appendix).

**Figure 2.**
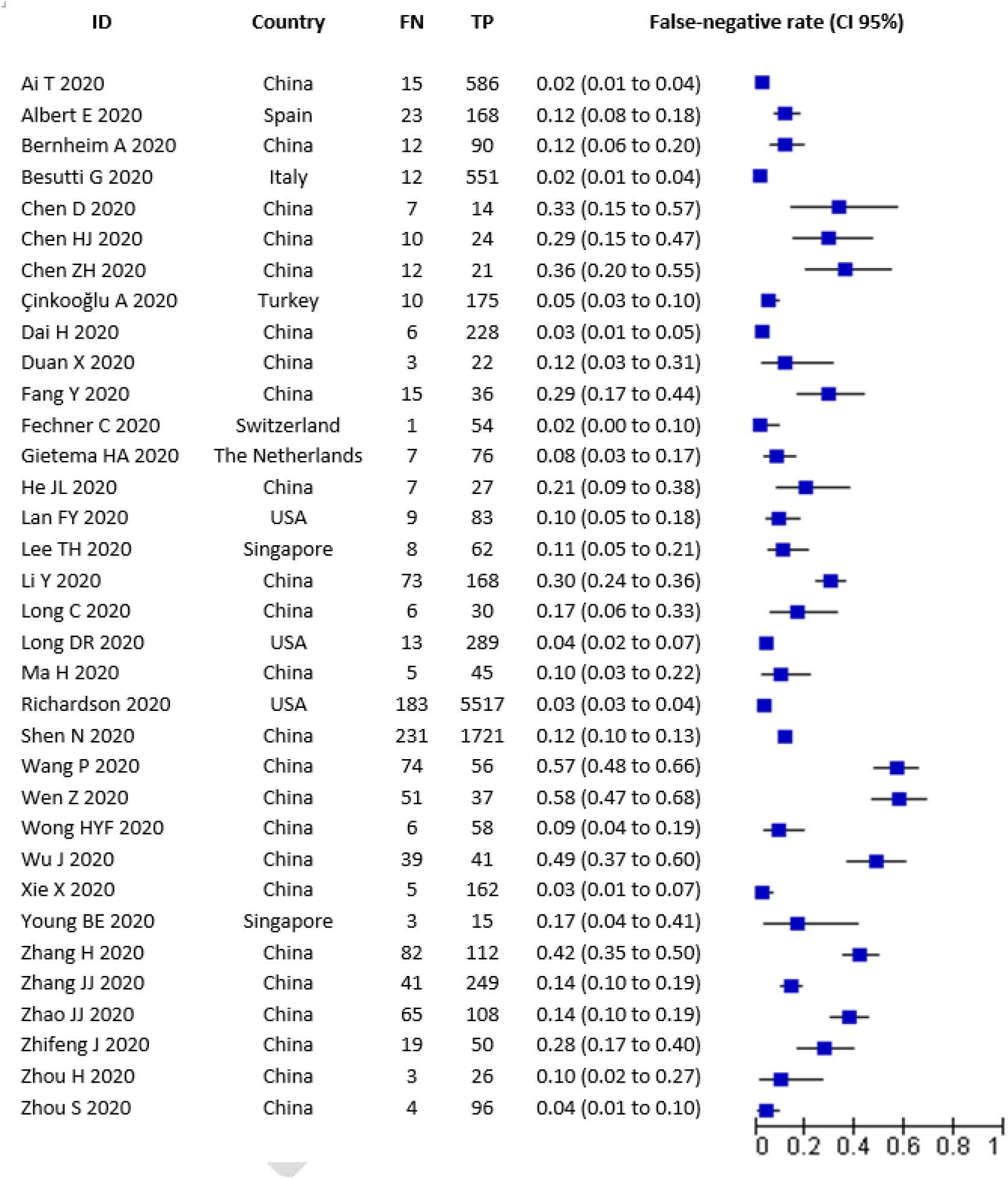
Forest plot included studies.

According to the QUADAS-2 tool, the domain most affected by a high risk of bias was the flow and timing domain, as some studies had not repeated the RT-PCR testing to all patients with negative results at initial testing (36, 44, 45, 51, 53, 54); besides, some studies did not provide information about the interval of time for the administration of a new RT-PCR assay. Regarding the patient selection domain, the risk of bias and applicability concerns were judged as high or unclear for several studies selecting patients assessed by RT-PCR plus Chest CT findings or serology tests. In most of the studies was unclear whether the administration of these tests was the standard protocol of management, or if the authors only enrolled patients undergoing all tests (33-35, 37-40, 46, 47, 50, 52, 56, 57, 59, 63, 66).

In regards to the index test domain, details about the criteria for positive results, such as the target genes under assessment of the SARS-CoV-2 nucleic acid detection kit used, were not provided by several studies. Their risk of bias and applicability were judged as unclear in both cases (33, 35-44, 46, 47, 49, 50, 52, 53, 56, 59, 63, 65, 66). Finally, two studies were judged as unclear in the reference standard domain, since the authors did not report in detail the characteristics of the repeated RT-PCR and their administration (38, 51). Six studies were considered as at low risk of bias in all QUADAS-II domains assessed (48, 55, 58, 60, 61, 64), while 20 were considered as at unclear risk due to at least one domain was judged with unclear risk of bias. The remaining eight studies were considered at high risk of bias (at least one domain judged with high risk) (36, 37, 45, 50, 51, 53, 54, 62).The analysis of limitations carried out with the adapted JBI case-series tool provided a similar assessment of the quality of included studies due to the uncertainty regarding the consecutive inclusion of patients and follow-up time after the first RT-PCR result. Additionally, due to the selection of patients, the majority of included studies were not an adequate sample of the target population (S4 Appendix).

### Findings

We analysed information from 34 studies collecting information from 12,057 patients confirmed to have SARS-CoV-2 infection and 1060 cases with RT-PCR negative findings in their initial assessment. False-negative rates ranged from 0.018 (44) to 0.58 (56) (Figure 2).

The summary estimate of the false-negative rate was 0.13 (95% CI 0.09 to 0.19). The data were characterised by a considerable between-study heterogeneity (tau-squared = 1.39). The 90% prediction interval ranged from 0.02 to 0.54.

Assessment of the effect of potential sources of heterogeneity was limited due to the lack of separate information of relevant subgroups (Table 2). There were no differences related to the duration of symptoms at the time of the first RT-PCR test based on information derived from nine studies provided means and medians of symptoms onset (Table 2). Comparison of false-negative rates of studies using different RT-PCR kits targetting (nucleocapside N-gene and/or ORF1ab gene) makes no significant differences (Table 2). In addition, most of the studies (28 out of 34) reported a mixture of specimens collected for RT-PCR assessment; those reporting the use of nasopharyngeal swabs provided a range of false-negative from 0.018 to 0.33, while those reporting the additional use of oropharyngeal swabs reported a range of false-negative from 0.02 to 0.33. Only the analysis by country (China versus other countries) showed a potential effect in the summary estimations; studies developed in other countries provide a false-negative pooled estimation of 0.06 (CI 95%= 0.04 to 0.09; 90% prediction interval 0.02 to 0.17; tau-squared= 0.36). Using meta-regression, we found a positive association of country with the false-negative rate (Table 2).

**Table 2.** Assessment of sources of heterogeneity.

| Variable |  | Number of studies (patients) | Heterogeneity (Tau-squared) | P-value |
| --- | --- | --- | --- | --- |
| Days of symptoms (average/median) | Less than 5 days | 3 (120) | 0.01 | 0.145 |
|  | Five days or more | 6 (817) | 0.87 |  |
| PCR target | N gene | 8 (2911) | 1.09 | 0.448 |
|  | No N gene | 5 (615) | 0.30 |  |
|  | ORF1ab gene | 10 (3188) | 0.91 | 0.144 |
|  | No ORF1ab gene | 3 (338) | 0.00 |  |
| Country | China | 24 (4798) | 1.31 | 0.002 |
|  | Other countries | 10 (7259) | 0.36 |  |
| Type of design | Accuracy | 12 (1798) | 1.52 | 0.407 |
|  | Cohorts/case series | 22 (10259) | 1.28 |  |
| Risk of bias | High risk | 8 (8947) | 0.79 | Reference |
|  | Unclear risk | 20 (2549) | 1.31 | 0.357 |
|  | Low risk | 6 (561) | 0.60 | 0.004 |

Additional post-hoc analysis by type of study did not provide a reduction of the observed heterogeneity (accuracy studies = 0.16, 95% CI 0.08 to 0.28, tau-squared= 1.52; cohorts/case-series=0.12, 95% CI 0.08 to 0.18, tau-square = 1.28). An analysis by the global risk of bias (based on the QUADAS-II domains) showed a difference between high risk versus low risk studies (high-risk studies = 0.08, 95% CI 0.04 to 0.14, tau-square = 0.79; low-risk studies=0.33, 95% CI 0.20 to 0.49, Tau-square =0.60), although the heterogeneity remains similar to those reported for the total group (Table 2).

Since we are not able to warrant that the summary estimate provided by the meta-analysis is a valid representation of the false-negative rate that can be expected in current practice, because of the very large heterogeneity, we instead used the estimated prediction interval in the analysis of the certainty of the evidence using the GRADE approach.

### Certainty of the evidence

We used the estimated prediction interval of the main meta-analysis to develop a summary of findings following the GRADE approach. We illustrated the consequences of the range of false-negative rates in a population of 100 tested, according to three different prevalence estimates seen in the current clinical practice for participant stakeholders and similar to those estimated by the included studies (10%, 30%, and 50%) (Figure 3). Using a prevalence of 50%, we found that 1 to 27 cases would be misdiagnosed and then they would not receive adequate clinical management; in addition, they could require repeated testing at some point in their hospitalisation or require another testing for competing diagnoses. The quality of the evidence was judged to be very low due to issues related to the risk of bias, indirectness, and inconsistency (Figure 3). This numerical approach should be interpreted with caution due to the multiple limitations of the evidence described above (Figure 3).

**Figure 3.**
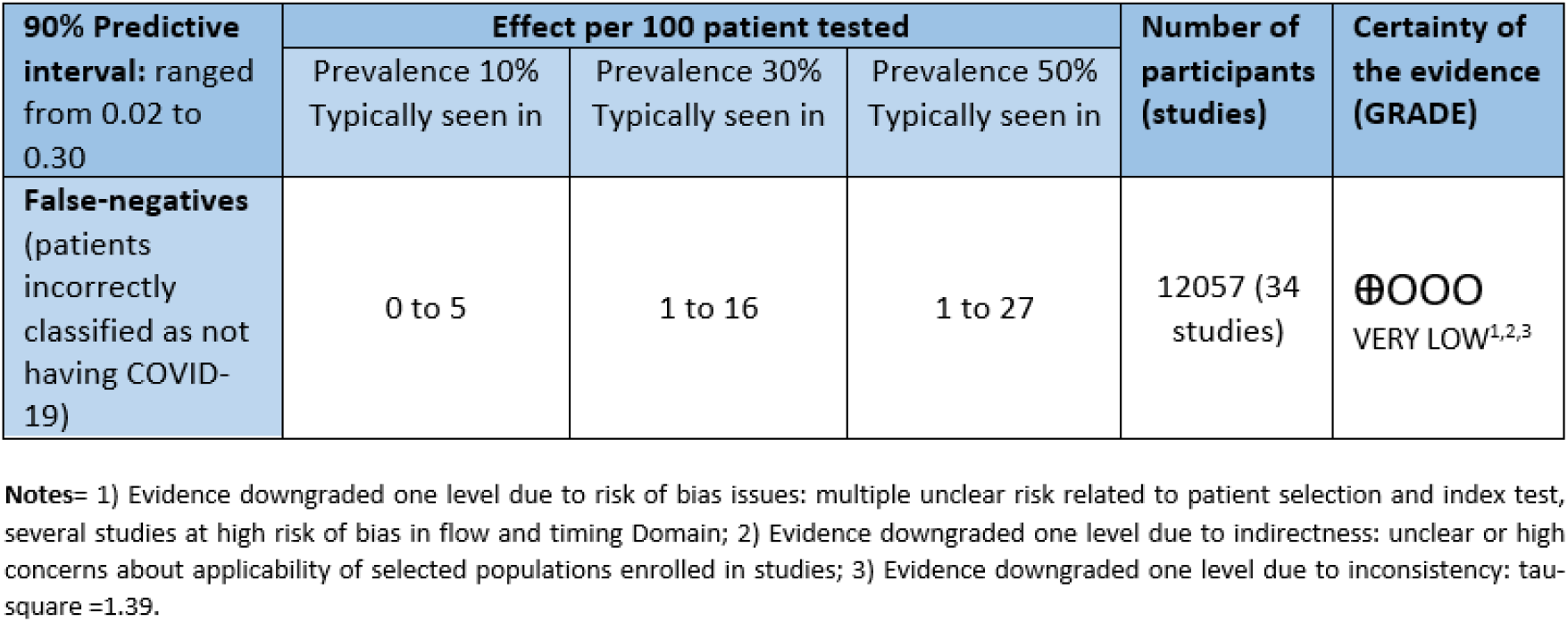
Certainty of the evidence (GRADE assessment)

## DISCUSSION

Our systematic review identified 34 studies and 12,507 participants providing information about the proportion of false-negative (FN) cases in the detection of SARS-CoV-2 by RT-PCR assays at first use. Individual studies estimates of false-negative rate ranged from 0.018 to 0.58. Included studies were affected by several sources of potential bias, especially related to the administration of an additional RT-PCR to rule in/rule out the presence of SARS-CoV-2 infection, the analysis of a selected sample of COVID-19 patients, as well as the unclear report of key index test characteristics.

The meta-analysis of the FN rates showed a considerable variability of data not explained by any of the foreseen potential sources of heterogeneity. This variability is a limitation for the interpretation of the mean proportion of the FN results as a summary estimate. Kucirka et al. also detected similar uncertainties in their Bayesian modelling of false-negative rates of RT-PCR by time since exposure, based on information from seven studies and 1330 respiratory samples (68). As an alternative, we chose to illustrate the impact of this heterogeneity by showing the number of false-negative cases expected in a cohort of 100 patients tested under three different prevalence of the disease scenarios. We based our calculations on the limits of the false-negative prediction interval. Using a prevalence of 50%, we found that up to 27 cases would be misdiagnosed and then they would not receive adequate clinical management. We emphasised that these numerical approaches should be interpreted with caution due to very low quality of evidence.

Our systematic review faced other challenges in its development. First, our study was initially planned as a rapid review aiming to provide a quick response to our local clinicians at the beginning of the COVID-19 pandemic. Due to the permanent involvement of clinicians managing COVID-19 patients at this point, we were able to define a review question that responds to a clinical inquiry relevant to current clinical practice (69-71). However, due to the increasing number of publications potentially eligible to answer the review question, our approach evolved into a living-systematic review with regular updates of the evidence. This manuscript reflects the third update of our literature searches with information current up to July 2020. To promote transparency in the development of this review, we have uploaded our previous results in the Open Science Framework repository for public consultation (https://osf.io/iserd/). We plan to perform additional searches after the publication of this manuscript to keep the conclusions as update as possible.

A second challenge is related to the type of studies providing information about the false-negative rate associated with RT-PCR at initial testing. We expected to find studies specifically aimed to estimate the number of initial negative results of RT-PCR assays, with further confirmation of SARS-CoV-2 infection with an additional RT-PCR within the following days to the first result. On the contrary, we found that the reporting of false-negative rate was not the primary aim of any of the include studies. In some cases, these figures were reported as descriptive statistics of the collected sample. Although we carried out a comprehensive and sensitive search strategy including major databases and repositories of preprint publications, we cannot discard that some eligible studies have not been identified yet due to the limitation of the reporting in key study sections, such as the abstract and methods.

Finally, as we have remarked in the findings section of this review, we found a considerable heterogeneity of data not explained by the statistical analysis performed. Suggested sources of heterogeneity such as the type of specimen collected, the time to onset of symptoms (as an approach to viral load), as well as the name of the RT-PCR kit used (to know essential characteristics as their analytical properties), were insufficiently reported or not reported at all for collected studies. This variability on COVID-19 testing data and the challenge to provide a pooled estimation with a useful clinical meaning have been previously remarked as the main constraint in the development of systematic reviews on this field (72). Despite our efforts in the analysis of data, we only were able to find some reduction of this variability comparing those studies performed in China versus those carried out in other countries (i.e. USA, Singapore, and the Netherlands). We believe that information provided by Chinese studies reflects early experiences with the diagnosis of COVID-19; their findings are probably affected by several unreported issues, such as the RT-PCR kits in use (likely the first kits developed for SARS-CoV-2 detection), the lack of standardised methods for COVID-19 testing and, in general, the limited knowledge about this new infection at the beginning of 2020.

Despite the heterogeneous information answering the review question, our study carried out a rigorous assessment of potential sources of bias, a formal statistical analysis of results and a final evaluation of the certainty of the evidence under a well-known system (GRADE). Although not all studies included in this review were accuracy studies, we decided to apply the QUADAS-II tool regardless of the type of design. However, even though QUADAS-II was not developed to evaluate case series, we preferred to standardise the quality assessment to report on a common pool of issues. We added as an appendix the assessment of all studies using an adapted checklist tool for case-series to provide complementary information to this assessment. Due to the multiple difficulties associated with the lack of reporting of included studies, and due to the high probability of new studies being published in the short-term, we provided some recommendations for future studies candidates to be included in an update of this review:

- Inclusion of a series of consecutive patients instead of selected groups, to avoid spectrum bias.
- Description of RT-PCR scheme in use, including target genes under assessment and positivity criteria.
- Description of preanalytical steps (conservation of samples, time until being sent to the laboratory, training of personal).
- Clear reporting of the time since the onset of symptoms, especially for those patients with clinical findings at admission
- Reporting of the number of additional RT-PCR assays performed
- Details about the application of the reference standard, including the time of administration after the index test (initial RT-PCR)
- If possible, database sharing could allow re-analyses by independent researchers, including individual-patient data (IPD)-meta-analysis and increasing thus the confidence on the new evidence
- Adding serological samples to a cohort of individuals with compatible symptoms and negative PCR to warrant an independent verification of infection.

## CONCLUSIONS

Our findings reinforce the need for repeated testing in patients with suspicion of being infected, due to either clinical or epidemiological reasons, given that up to 54% of COVID-19 patients may have an initial negative RT-PCR result (certainty of evidence: very low). The collected evidence has several limitations in terms of risk of bias and applicability; besides, lack of reporting of several key factors remains a significant constraint for a comprehensive analysis of collected data. A new update of this review when additional studies become available is warranted.

## Data Availability

The study protocol is available online at https://osf.io/jserd/. Most included studies are publically available. Additional data are available upon reasonable request.

**LIST OF ABBREVIATIONS**
Chest CT: Chest Computed tomography
COVID-19: coronavirus disease 2019
GRADE: Grading of Recommendations, Assessment, Development and Evaluation
PRISMA: Preferred Reporting Items for Systematic Reviews and Meta-Analyses
QUADAS-II: Quality Assessment of Diagnostic Accuracy Studies-II tool
RT-PCR: reverse transcription-polymerase chain reaction
SARS-CoV-2: Severe Acute Respiratory Coronavirus 2
WHO: World Health Organization

## DECLARATIONS

### Ethics approval and consent to participate

Not applicable

### Consent for publication

Not applicable

### Availability of data and material

The datasets used and/or analysed during the current study are available from the corresponding author on reasonable request. The study protocol is available online at https://tinvurl.com/vvbgqya.

### Competing interests

The authors declare that they have no competing interests

### Funding

No specific funding was received for the development of this research

### Authors’ contribution

IAR, DBG, DS and JZ conceived the study. IAR, DBG, DSR, RDC, JAPM and JZ designed the study. IAR, DBG, DSR, PZA screened titles and abstracts for inclusion. IAR, DBG, DSR, PZA, AR and JZ extracted and analysed data. RDC, JAPM, AC, OS and NL assisted in the interpretation from a clinical viewpoint. IAR, DBG, DSR and JZ wrote the first draft, which all authors revised for critical content. All authors approved the final manuscript. IAR and JZ are the guarantors. The corresponding author attests that all listed authors meet authorship criteria and that no others meeting the criteria have been omitted.

## Acknowledgements

Ingrid Arevalo-Rodriguez is funded by the Instituto de Salud Carlos III through the “Acción Estratégica en Salud 2013-2016 / Contratos Sara Borrell convocatoria 2017/CD17/00219” (Co-funded by European Social Fund 2014-2020, “Investing in your future”).

## Notes

### Competing Interest Statement

The authors have declared no competing interest.

### Clinical Protocols

https://osf.io/jserd/

### Funding Statement

No external funding received

### Summary of Updates

Last search performed on July 17-2020. Findings updated

